# The association between proportion of night shifts and musculoskeletal pain and headaches in nurses: a cross-sectional study

**DOI:** 10.1101/2023.02.09.23285689

**Authors:** Jon Are Stavås, Kristian Bernhard Nilsen, Dagfinn Matre

## Abstract

**Objectives:** To investigate whether a higher proportion of night shifts is associated with a higher occurrence of musculoskeletal pain and headaches. Furthermore, to investigate whether sleep duration can mediate this potential association.

**Method:** The study included 684 nurses in rotating shift work (day, evening, night) who responded to a daily questionnaire about working hours, sleep, and pain for 28 consecutive days. The data were treated as cross-sectional data.

**Results:** A negative binomial regression analysis adjusted for age and BMI revealed that working a higher proportion of night shifts is not associated with a higher occurrence of musculoskeletal pain and headaches. On the contrary, those working ≥ 50 % night shifts had a significantly lower occurrence of pain in the lower extremities than those who worked <25 % night shifts (IRR 0.69 95 % CI 0.51, 0.94). There was no indication of a mediation effect with total sleep time (TST).

**Conclusion:** The results of this study indicate that working a higher monthly proportion of night shifts is not associated with a higher occurrence of musculoskeletal pain and headaches.

**What is already known on this topic?:** A few studies have investigated the association between proportion of night shifts and musculoskeletal pain and headaches, but the findings are conflicting, and the studies are largely based on retrospective questionnaires.

**What this study adds:** This study gives further knowledge about the association between proportion of night shifts and musculoskeletal pain and headaches.

**How this study might affect research, practice or policy:** The results have implications for the design of future studies on the effect of shift work on musculoskeletal pain and headache. The negative effect of night shifts are not necessarily dose-dependent, and real-life studies must seek to account for the “healthy worker effect”.

## INTRODUCTION

Shift work is associated with musculoskeletal pain and headaches (1, 2), but little is known about how the intensity of shift work exposure is related to musculoskeletal pain and headaches. An increasingly large proportion of the population is working shift (3). There is, need for a better understanding of the association between shift work and its associated burdens.

Musculoskeletal pain and headaches have major negative consequences for both the individual and society (4, 5). According to the Global Burden of Disease study, low back pain and headache disorders are the two leading causes of years of healthy life lost due to disability (YLD) worldwide (6). Consequently, musculoskeletal pain and headaches impose vast financial and individual costs.

Since shift work is necessary in multiple occupational sectors and industries, it is imperative to determine which characteristics of shift work that are associated with the occurrence of musculoskeletal pain and headaches. Shift work which includes night shifts, may pose a greater health risk than shift work without nights. In a previous study, we found that night shifts was associated with pain complaints the following day in nurses (7), and that headache tended to increase from the second to the third night shift (8). Reduced sleep time and disturbances in the circadian rhythm may contribute to this association (9). Shift workers who work night shifts have an increased risk of experiencing reduced sleep length and sleep disturbances (10), and reduced sleep length and sleep disturbances are associated with musculoskeletal pain and headaches (11-13). Hence, reduced sleep length may potentially act as a mediator between night shifts and musculoskeletal pain and headaches. However, it is unclear whether a higher proportion of night shifts per unit of time is associated with musculoskeletal pain and headaches. These knowledge gaps may be approached by investigating whether a higher proportion of night shifts are associated with a higher occurrence of musculoskeletal pain and headaches, and whether sleep duration is a part of this putative association.

The proportion of night shifts is a working time pattern relevant for health, as described by Härmä et al. (14). In the present study we investigate whether a higher monthly proportion of night shifts are associated with a higher occurrence of musculoskeletal pain and/or headaches. A few studies have investigated this association, but the findings are conflicting, and the studies are largely based on retrospective questionnaires, which may be affected by reporting bias (15-23).

The objective of this study was to investigate the association between night shift frequency, musculoskeletal pain, and headaches. We hypothesized that a higher proportion of night shifts would be associated with a higher occurrence of musculoskeletal pain and headaches. We also hypothesized if sleep duration could mediate this association.

## METHODS

### Design and study population

The data is from a diary study carried out by The National Institute of Occupational Health in Norway (STAMI) through the project “Shift work, sleep and pain” from October 2014 to November 2015.

An invitation was sent by post or by e-mail to a random sample of members of the Norwegian Nurses Association (NSF) (n=22,500), to reduce selection bias. The inclusion criteria were that the participants had to be authorized nurses, working in more than 50% position, being between 18 and 63 years of age, and be working in a 2-shift (day, evening) or 3-shift (day, evening, night) schedule or night shifts only. Participants who were pregnant, breastfeeding, or had been on sick leave for more than two weeks in the last six months were excluded. A total of 5,400 participants received a further invitation to participate in the study. 1032 participants answered the questionnaire at baseline. Of these, 727 participants also answered a diary questionnaire for up to 28 days. After correcting for the inclusion criteria, 684 nurses completed both the questionnaire at baseline at the diary questionnaire for a minimum of seven days.

A priori power calculations assuming 80% power indicated that a sample size of 100 was sufficient to detect a 20% difference in pain complaints, if pain complaints were treated as a continuous variable. For the present analysis, the pain variable was dichotomized, which reduces statistical power. To compensate, the number of participants was increased 6-7 fold.

### Compliance with ethical standards

All participants gave written informed consent to participate in the study, which was conducted in accordance with the Declaration of Helsinki and was approved by the Norwegian Regional Committee for Medical Research Ethics (approval number 2012/199/REK sør-øst B).

### Data collection

The questionnaire at baseline contained questions about demographic conditions, lifestyle, sleep, physical and mental complaints as well as psychosocial and physical working conditions. Participants who answered the questionnaire were included in the study. After inclusion, an SMS (or e-mail if not owning a smartphone) was sent to the participants daily at 21.00 with a reminder to answer a diary questionnaire as soon as possible. Recall bias is a known limitation in questionnaire studies. To limit recall bias, the participants were asked to answer daily questions about pain, working hours, and sleep for a total of 28 consecutive days. Answers received 24 hours or more after the reminder, were not considered.

### Exposure variables

Morning shift was defined as a shift that started between 05:00 and 12:00. An evening shift was defined as a shift that started between 12:01 and 18:00, while a night shift was defined as a shift that started between 18:01 and 04:59. Hence, each workday was categorized accordingly. Days without work was categorized as days off.

The number of morning, evening, and night shifts worked for each participant for 28 days was calculated by aggregating the variables. The proportion of night shifts was then calculated by dividing the number of night shifts by all shifts worked. The proportion of night shifts was categorized as an ordinal variable. The categories used were <25%, 25-49.9%, and ≥ 50%, as suggested by Härmä et al. (14).

### Sleep as a potential mediator

The diary sleep questions were derived from Carney et al. (24). These questions were:

1. “What time did you get into bed?”, 2. “What time did you try to fall asleep?”, 3. “How long time (in hours and minutes) did it take for you to fall asleep?” which is described as sleep onset latency (SOL), 4. “How many times did you wake up, not counting your final awakening?”, 5. “In total, how long did these awakenings last?” which is described as wake after sleep onset (WASO), 6. “What time was your final awakening?”, 7. “What time did you get out of bed after your main sleep?”, 8. “Did you wake up earlier than planned?”, 9. “How long did you sleep in addition to your main sleep during the past 24 h?”

Daily total sleep time (TST) was calculated by subtracting SOL and WASO from the time difference between when the participants was trying to sleep and the time of the final awakening. TST for all days was then summed for each participant. Furthermore, the variable was aggregated to find the number of days responded. The total sum of the TST was finally divided by the number of days responded to find the average TST. The average TST was treated as a continuous variable.

### Outcome variables

Self-reported subjective musculoskeletal pain complaints and headache were measured using a diary questionnaire in which the participants scored pain in the last 24 hours on a four-point Likert scale. The options were not troubled by pain=0, somewhat troubled by pain=1, fairly troubled by pain=2, and very troubled by pain=3. Participants rated pain in five regions: the neck, shoulder, and upper back, lower back, upper extremities, lower extremities, and headaches.

In this study we wanted to study the overall effect of night shifts on pain prevalence, and not the variation in pain intensity. Thus, for each day and pain region, the pain ratings were dichotomized and summed for the data collection period of 28 days to find the number of days with pain. The five outcome variables, musculoskeletal pain and headache, were treated as discrete count variables.

### Statistical analysis

All analyzes were done in R (RStudio Team, 2022). The estimates from the analyzes were presented with a 95% confidence interval (CI). The significance level was set at ≤ 0.05. The analysis was adjusted for age and BMI.

In this study, the data collected over 28 days were treated as cross-sectional data. Negative binomial regression was used, as the model was overdispersed (25), to examine the association between the proportion of night shifts and the outcome variables. The reference category was <25% night shifts, which was compared with 25-49.9% and ≥ 50% night shifts. Furthermore, it was investigated if TST could mediate the association between the proportion of night shifts and the outcome variables. If the combined effect for path a and path b is significant, it can be concluded that there is a mediation (see figure 1) (26). The indirect effect can be found by multiplying the partial derivative of the equation for path a with respect to X by the partial derivative of the equation for path b and path c’ with respect to M (27). Linear regression was used to analyze path a, and negative binomial regression was used for path b and path c’. As described in Geldhof, Anthony (27), when linear regression has been used in path a and negative binomial regression in path b and path c’, the conditional indirect effect can be estimated by the formula:

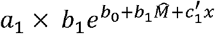

**Figure 1.**
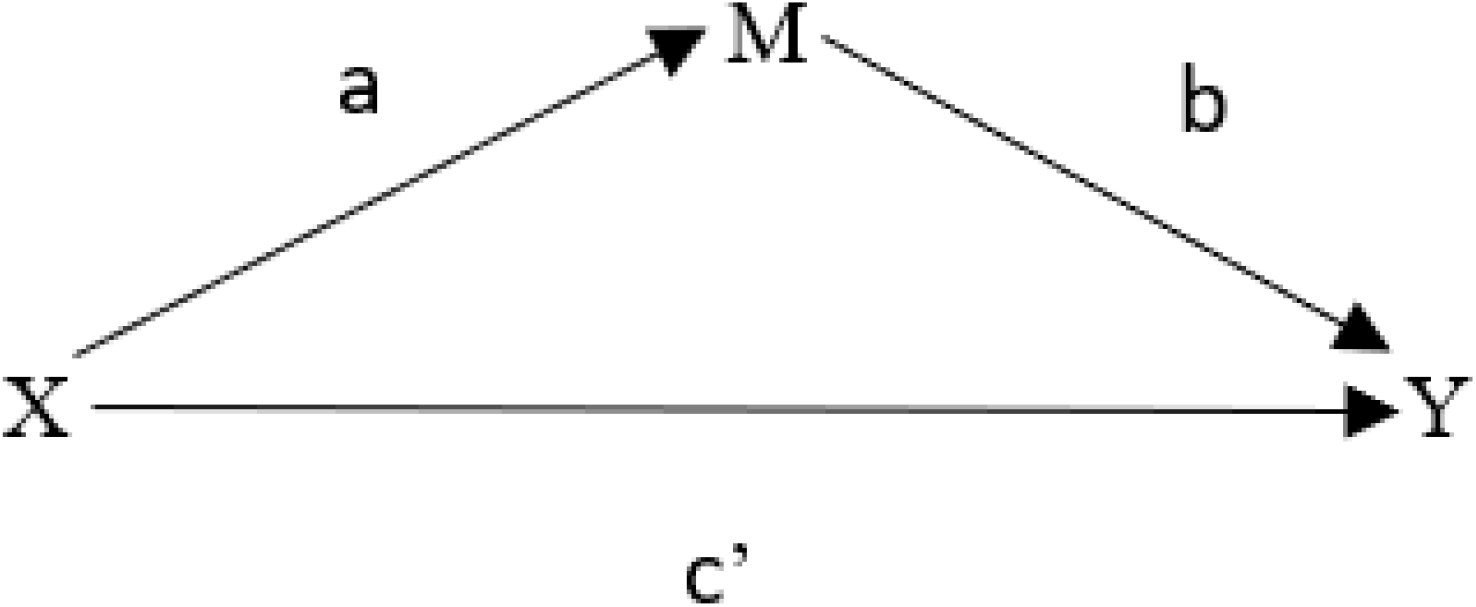
Mediation model. X is the exposure variable, M is the mediator, and Y is the outcome variable. The indirect effect is the combined effect between path a (X➔M) and path b (M➔Y). The direct effect is path c’ (X➔Y).

To calculate the confidence intervals for the conditional indirect effect, bootstrap was used as proposed in Geldhof, Anthony (27). 1000 replicates were calculated using bootstrap. In all analysis adjustment were made for age and body mass index (BMI) as the variables theoretically can be associated with both the exposure and outcome (28-31).

In addition, an offset variable was used for all the unadjusted an adjusted analyzes with the outcome variables to adjust for the difference in number of days responded to the diary questionnaire. Offset variables can be used in negative binomial regression to adjust for differences in time for count variables (25).

## RESULTS

### Descriptive statistics

The sample consisted of 684 nurses in shift work varying from nurses who worked a 2-or 3-shift schedule or night shift only. The diary questionnaire was answered 84-85% of the days. Of the 727 participants who responded to diary questionnaire, 43 participants were excluded due to the following reasons: they had responded less than seven days to the questionnaire (n=22), not working as a nurse (n=14), were on sick leave (n=1), or had missing data for one of the confounding variables (n=6). Table 1 presents descriptive statistics consisting of demographic variables, work related variables, pain, and sleep. Mean age was 40.9 ± 11.1 years with a range of 22-63 years. Table 2 presents descriptive statistics for total sleep time (TST) for the different exposure groups used in the analysis.

**Table 1.** Descriptive statistics with demographic variables, work related variables, pain, and sleep.

| **Variables** | **Participants (n=684)** |
| --- | --- |
| Sex (female), n (%) | 614 (90) |
| Age, year, mean ± SD | $40.9 \pm 11.1$ |
| Body mass index, kg/m<sup>2</sup>, mean ± SD | $24.7 \pm 4.1$ |
| Percentage of full-time position (%) |  |
| 50-75 % | 21.6 |
| 76-90 % | 19.7 |
| >90 % | 58.0 |
| Naps on the night shifts, n (%)<sup>b</sup> | 224 (34) |
| Morning shift, %<sup>c</sup>, median (IQR) | 52.9 (37.5–63.6) |
| Evening shift, %<sup>c</sup>, median (IQR) | 25.5 (14.3–35.9) |
| Night shift, %<sup>c</sup>, median (IQR) | 17.6 (6.7–33.3) |
| Neck-, shoulder-, and upper back pain, number of days, median (IQR) | 4 (1–12) |
| Low back pain, number of days, median (IQR) | 2 (0–7) |
| Pain in the upper extremities, number of days, median (IQR) | 0 (0–3) |
| Pain in the lower extremities, number of days, median (IQR) | 2 (0–9) |
| Headache, number of days, median (IQR) | 4 (1–8) |
| TST, hours, mean ± SD | 6.7 ± 1.0<sup>a</sup> |
| <sup>a</sup>n=645 |  |
| <sup>b</sup>occasionally or more often |  |
| <sup>c</sup>proportion of shifts in percent out of all shifts |  |
| SD: Standard deviation. IQR: Interquartile range. TST: Total sleep time. |  |

**Table 2.** Total sleep time (TST) in hours for different proportion of night shifts.

| Proportion of night shifts (%) | TST, hours, mean ± SD |
| --- | --- |
| <25 | 6.7 ± 0.9 |
| 25-49.9 | 6.7 ± 0.7 |
| ≥50 | 6.5 ± 1.0 |

### Proportion of night shifts and musculoskeletal pain and headaches

Those who worked ≥ 50 % night shifts had significantly lower occurrence of pain in the lower extremities than those who worked <25 % night shifts in the adjusted analysis (IRR 0.69 95 % CI 0.51, 0.94) (Table 3). For pain in other regions, no significant association was found with proportion of night shifts. As a sensitivity analysis, only nurses working a 3-shift schedule were included in the analysis. No significant associations were found in the sensitivity analysis (Table 4).

**Table 3.**
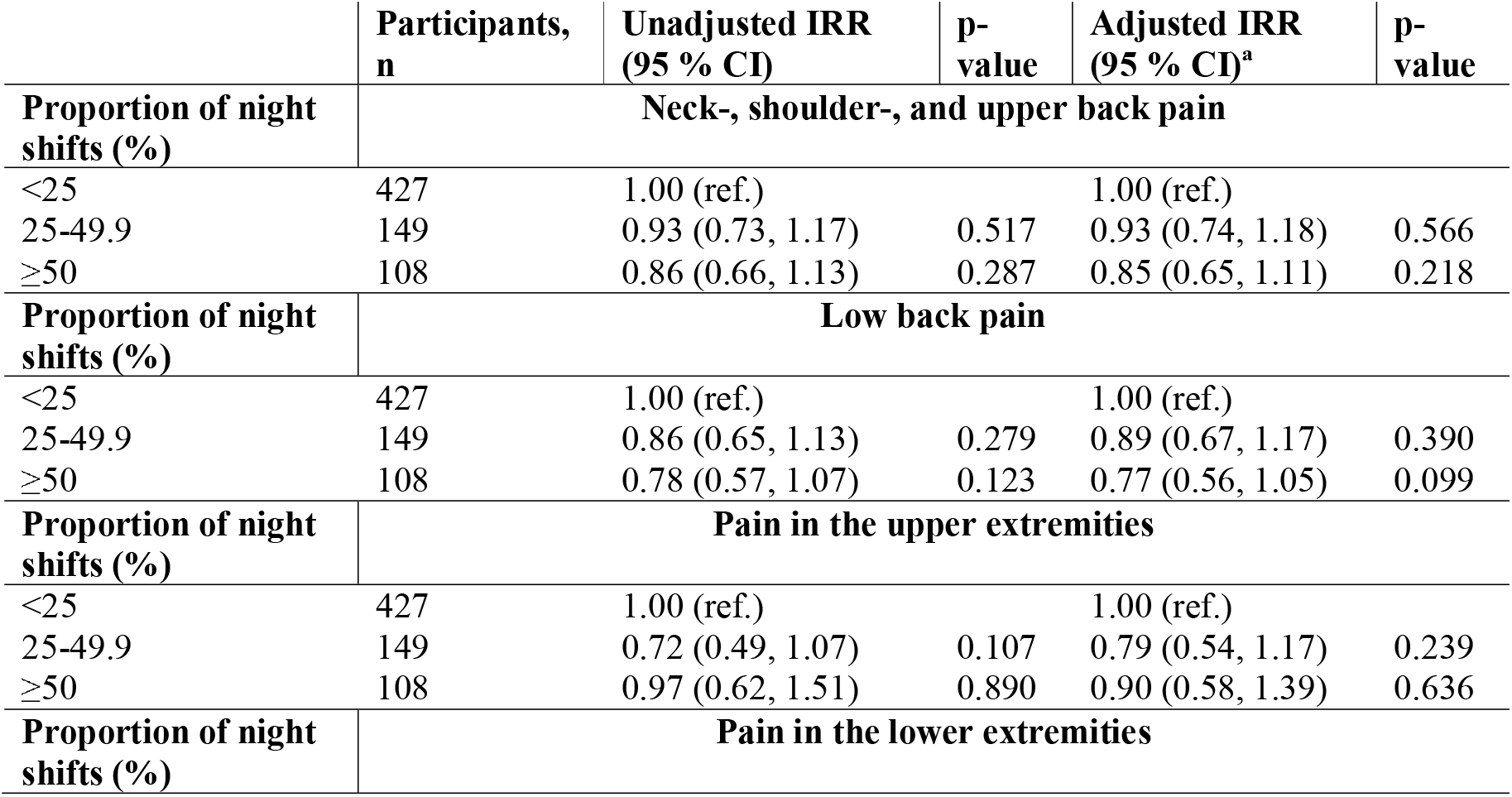
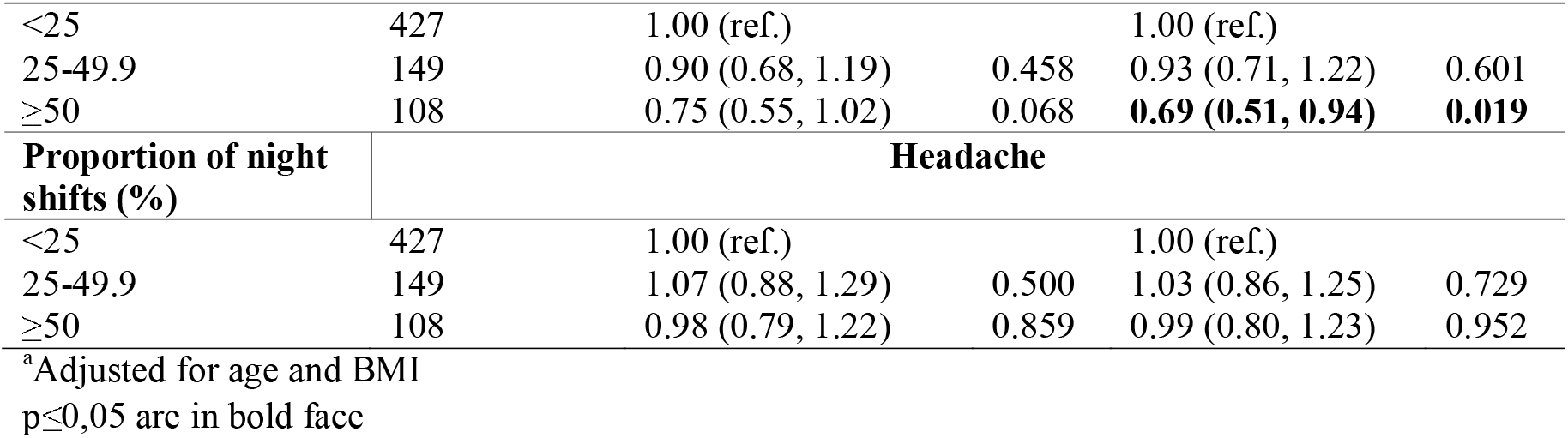
Association between proportion of night shifts and musculoskeletal pain and headaches among nurses working a 2-, 3-shift schedule or night shifts only (n=684). Incidence rate ratio (IRR) with 95 % confidence intervals (CI) and p-values from negative binomial regression analysis.

|  | Participants, n | Unadjusted IRR (95 % CI) | p-value | Adjusted IRR (95 % CI)<sup>a</sup> | p-value |
| --- | --- | --- | --- | --- | --- |
| **Proportion of night shifts (%)** | **Neck-, shoulder-, and upper back pain** |  |  |  |  |
| <25 | 427 | 1.00 (ref.) |  | 1.00 (ref.) |  |
| 25-49.9 | 149 | 0.93 (0.73, 1.17) | 0.517 | 0.93 (0.74, 1.18) | 0.566 |
| ≥50 | 108 | 0.86 (0.66, 1.13) | 0.287 | 0.85 (0.65, 1.11) | 0.218 |
| **Proportion of night shifts (%)** | **Low back pain** |  |  |  |  |
| <25 | 427 | 1.00 (ref.) |  | 1.00 (ref.) |  |
| 25-49.9 | 149 | 0.86 (0.65, 1.13) | 0.279 | 0.89 (0.67, 1.17) | 0.390 |
| ≥50 | 108 | 0.78 (0.57, 1.07) | 0.123 | 0.77 (0.56, 1.05) | 0.099 |
| **Proportion of night shifts (%)** | **Pain in the upper extremities** |  |  |  |  |
| <25 | 427 | 1.00 (ref.) |  | 1.00 (ref.) |  |
| 25-49.9 | 149 | 0.72 (0.49, 1.07) | 0.107 | 0.79 (0.54, 1.17) | 0.239 |
| ≥50 | 108 | 0.97 (0.62, 1.51) | 0.890 | 0.90 (0.58, 1.39) | 0.636 |
| **Proportion of night shifts (%)** | **Pain in the lower extremities** |  |  |  |  |
| <25 | 427 | 1.00 (ref.) |  | 1.00 (ref.) |  |
| 25-49.9 | 149 | 0.90 (0.68, 1.19) | 0.458 | 0.93 (0.71, 1.22) | 0.601 |
| ≥50 | 108 | 0.75 (0.55, 1.02) | 0.068 | **0.69 (0.51, 0.94)** | **0.019** |
| **Proportion of night shifts (%)** | **Headache** |  |  |  |  |
| <25 | 427 | 1.00 (ref.) |  | 1.00 (ref.) |  |
| 25-49.9 | 149 | 1.07 (0.88, 1.29) | 0.500 | 1.03 (0.86, 1.25) | 0.729 |
| ≥50 | 108 | 0.98 (0.79, 1.22) | 0.859 | 0.99 (0.80, 1.23) | 0.952 |

<sup>a</sup>Adjusted for age and BMI p≤0,05 are in bold face

**Table 4.** Association between proportion of night shifts and musculoskeletal pain and headaches among nurses working 3-shift schedule (n=551). Incidence rate ratio (IRR) with 95 % confidence intervals (CI) and p-values from negative binomial regression analysis.

|  | **Participants, n** | **Unadjusted IRR (95 % CI)** | **p-value** | **Adjusted IRR (95 % CI)<sup>a</sup>** | **p-value** |
| --- | --- | --- | --- | --- | --- |
| **Proportion of night shifts (%)** | **Neck-, shoulder-, and upper back pain** |  |  |  |  |
| <25 | 366 | 1.00 (ref.) |  | 1.00 (ref.) |  |
| 25-49.9 | 144 | 0.96 (0.75, 1.23) | 0.736 | 0.96 (0.75, 1.23) | 0.741 |
| ≥50 | 41 | 0.97 (0.64, 1.46) | 0.872 | 0.96 (0.64, 1.46) | 0.861 |
| **Proportion of night shifts (%)** | **Low back pain** |  |  |  |  |
| <25 | 366 | 1.00 (ref.) |  | 1.00 (ref.) |  |
| 25-49.9 | 144 | 0.85 (0.64, 1.13) | 0.265 | 0.87 (0.65, 1.16) | 0.349 |
| ≥50 | 41 | 0.74 (0.46, 1.20) | 0.225 | 0.74 (0.45, 1.19) | 0.211 |
| **Proportion of night shifts (%)** | **Pain in the upper extremities** |  |  |  |  |
| <25 | 366 | 1.00 (ref.) |  | 1.00 (ref.) |  |
| 25-49.9 | 144 | 0.71 (0.47, 1.08) | 0.111 | 0.77 (0.51, 1.15) | 0.204 |
| ≥50 | 41 | 0.67 (0.33, 1.35) | 0.265 | 0.64 (0.32, 1.27) | 0.202 |
| **Proportion of night shifts (%)** | **Pain in the lower extremities** |  |  |  |  |
| <25 | 366 | 1.00 (ref.) |  | 1.00 (ref.) |  |
| 25-49.9 | 144 | 0.92 (0.69, 1.22) | 0.571 | 0.94 (0.71, 1.24) | 0.660 |
| ≥50 | 41 | 0.74 (0.46, 1.19) | 0.215 | 0.70 (0.44, 1.13) | 0.143 |
| **Proportion of night shifts (%)** | **Headache** |  |  |  |  |
| <25 | 366 | 1.00 (ref.) |  | 1.00 (ref.) |  |
| 25-49.9 | 144 | 1.07 (0.87, 1.30) | 0.533 | 1.03 (0.85, 1.26) | 0.747 |
| ≥50 | 41 | 1.02 (0.73, 1.42) | 0.902 | 1.02 (0.73, 1.41) | 0.927 |

<sup>a</sup>Adjusted for age and BMI

### Mediation analysis

No indication of a mediation effect with TST as mediator between proportion of night shifts and for any of the pain regions was found (Supplementary table 1).

## DISCUSSION

The aim of the present study was to investigate whether the proportion of monthly night shifts in a was associated with the occurrence of musculoskeletal pain and headaches, and if total sleep time (TST) could mediate this potential association. We found that working a higher proportion of night shifts is not associated with a higher occurrence of musculoskeletal pain and headaches. On the contrary, those working ≥ 50 % night shifts had a significantly lower occurrence of pain in the lower extremities than those who worked <25 % night shifts. It was not found that TST could mediate the potential association between proportion of night shifts and musculoskeletal pain and headaches.

Previous studies that have examined the association between the proportion of night shifts and musculoskeletal pain and headaches have shown conflicting findings, with some studies finding an association and some not (15-23). Several explanations may contribute to the conflicting findings. Firstly, the previous studies are retrospective with a potential for reporting bias (32). Secondly, no distinction was made between those who had pain lasting only a few days and those who had pain every day over a longer period up to a year. This may have contributed to a higher number of cases classified with pain which possibly could bias the results towards the null hypothesis. Finally, the mentioned studies had different definitions for musculoskeletal pain and headaches. E.g., in the study by d’Ettorre et al. (15) work-related acute low back pain was defined as ‘activity-limiting lower back pain’ with a minimum duration of one day within 7 or 28 days. There was also a requirement that acute pain should have occurred at work. In June and Cho (16), low back pain was defined as ‘pain, aching, or stiffness’ at least once a month for one year. The different case definitions may have contributed to the different findings, depending on how strict the case definition was.

The support for a seemingly protective effect of working ≥ 50 % night shifts on lower extremity pain, compared to working <25 % night shifts, could potentially be explained by the so-called “healthy worker effect”. This bias assumes that individuals with health problems, or those who are most negatively affected by the exposure, quit their jobs, or are reassigned earlier (33). The same effect may explain why there was no significant association between the proportion of night shifts and pain in other musculoskeletal regions and headaches in this study. An example of “healthy worker effect” was seen in a prospective study of shift workers and the risk of being depressed, where those who were depressed at baseline had a greater risk of switching from shift work to only day work (34).

Mediation analysis with TST were carried out to better understand associations between proportion of night shifts and musculoskeletal pain and headache, despite no significant total effect was found for most of the analysis. According to Hayes and Rockwood (26), there can still be a significant indirect effect or mediation, even if there is no significant total effect. However, we did not find support for TST mediating the association between the proportion of night shifts and musculoskeletal pain and headaches. This is in contrast with a previous microlongitudinal study, from the same data material. In our previous study we found that TST could mediate the association between night shifts and lower extremity pain and abdominal pain, but this was not the case for other musculoskeletal pain and headaches (7). The difference between the present study and our previous study by Katsifaraki et al. (7) is that in the latter study, both TST and pain were analyzed from day to day, while in the present study TST and pain were respectively analyzed as mean and number of days over several weeks. In addition, Katsifaraki et al. investigated an increase in pain on a Likert-type scale from day to day, whereas in the present study pain was dichotomized each day as no pain or pain. Another study showed increased pain sensitivity the day after a night shift, but this was normalized after a night of normal sleep (35). It is therefore conceivable that there could be an association when examining TST and an increase in pain from day to day, but not when analyzing TST and number of days with pain. Kecklund and Axelsson (9) also argued that it was uncertain whether shift workers generally experience chronic sleep loss compared to day workers. In the present study several of the participants also answered that they occasionally or more often had naps on the night shifts. This could possibly influence the association, making those working night shift less prone to musculoskeletal pain and headaches. This type of pain-reducing effect of naps during night shifts has been reported by other (36).

### Strength and limitations

Strengths of the present study are a large, randomized sample of nurses and the longitudinal data collection. The data collection takes place close to exposure and outcome. It is conceivable that this can reduce reporting bias which can often be the case with questionnaires assessing complaints over weeks or months retrospectively (32).

Another strength of this study is that sensitivity analysis was carried out, analyzing only the participants who worked in a 3-shifts schedule. The sensitivity analysis produced essentially similar results, confirming no association between a higher proportion of night shifts and musculoskeletal pain and headaches.

Musculoskeletal pain and headaches were measured with a four-point Likert scale, which is a type of unidimensional NRS scale that is valid and reliable (37). The participants had a deadline of 24 hours to answer the diary questionnaire. Additionally, in this study, the cut-off value for pain was set to a minimum of “somewhat troubled”, and the variable was categorized as no pain or pain. Therefore, this study only distinguished between no pain and pain each day, making it difficult to compare the prevalence of pain to similar studies with a retrospective design. However, it is a strength that the sample was based on a randomized drawing from the Norwegian Nurses Association (NSF) and 90 % of the participant was women, which is similar to the nurse population in Norway (38), supporting the generalizability of this study.

A diary is preferable to a retrospective questionnaire, as sleep can vary greatly from night to night. Diaries have also been considered the gold standard for subjective sleep reporting (39). It is recommended to answer sleep diary questionnaire within one hour after the end of a sleep period (24). However, in the present study subjects were asked to report pain and sleep the last 24 hours. The sleep measures in this study may therefore have been prone to bias. It is also a limitation that we asked about exposure (night work) and outcome (pain) in the same questionnaire.

A limitation of this study is that the number of days data was collected from the diary questionnaire varied between participants from seven to 28 days. The data collection period had a duration of 28 days with a diary questionnaire, but the number of participants who completed the questionnaire became fewer towards the end of the study. Generally, excluding participants with missing data who may increase the risk of selection bias, and including them may increase the risk of information bias. To adjust for different number of days reported in the diary, an offset variable was used, which was the natural logarithm of the number of days answered on the diary questionnaire. This has also been done in other studies to adjust for time for follow-up (40). Thus, it is the rate of days with pain and not the number of days with pain that is analyzed (25).

### Concluding remarks

In this study, a higher monthly proportion of night shifts was not associated with a higher occurrence of musculoskeletal pain and headaches. On the contrary, a lower occurrence of pain in the lower extremities was found for those who worked ≥ 50 % night shifts compared to those who worked <25 % night shifts. The results have implications for the design of future studies on the effect of shift work on musculoskeletal pain and headache.

## Supporting information

STROBE checklist

## Data Availability

All data produced in the present study are available upon reasonable request to the authors

## Conflicts of interest

The authors declare no conflicts of interest.

## Funding

The study was funded by the National Institute of Occupational Health, Oslo, Norway.

## Notes

### Competing Interest Statement

The authors have declared no competing interest.

### Funding Statement

This study was funded by the National Institute of Occupational Health, Norway

### Author Declarations

The study was approved by the Norwegian Regional Committee for Medical Research Ethics (approval number 2012/199/REK sor-ost B).

## REFERENCES

1. Appel AM, Török E, Jensen MA, et al. The longitudinal association between shift work and headache: results from the Danish PRISME cohort. Int Arch Occup Environ Health. 2020;93(5):601–10.

2. Chang WP, Peng YX. Differences between fixed day shift nurses and rotating and irregular shift nurses in work-related musculoskeletal disorders: A literature review and meta-analysis. J Occup Health. 2021;63(1):e12208.

3. Eurofound. Sixth European Working Conditions Survey – Overview report (2017 update). 2017.

4. Briggs AM, Cross MJ, Hoy DG, et al. Musculoskeletal Health Conditions Represent a Global Threat to Healthy Aging: A Report for the 2015 World Health Organization World Report on Ageing and Health. The Gerontologist. 2016;56(Suppl_2):S243–S55.

5. Saylor D, Steiner TJ. The Global Burden of Headache. Semin Neurol. 2018;38(2):182–90.

6. James SL, Abate D, Abate KH, et al. Global, regional, and national incidence, prevalence, and years lived with disability for 354 diseases and injuries for 195 countries and territories, 1990–2017: a systematic analysis for the Global Burden of Disease Study 2017. The Lancet. 2018;392(10159):1789–858.

7. Katsifaraki M, Nilsen KB, Christensen JO, et al. Sleep duration mediates abdominal and lower-extremity pain after night work in nurses. Int Arch Occup Environ Health. 2019;92(3):415–22.

8. Katsifaraki M, Nilsen KB, Christensen JO, et al. Pain complaints after consecutive nights and quick returns in Norwegian nurses working three-shift rotation: an observational study. BMJ Open. 2020;10(9):e035533.

9. Kecklund G, Axelsson J. Health consequences of shift work and insufficient sleep. BMJ. 2016;355:i5210.

10. Fatima Y, Bucks RS, Mamun AA, et al. Sleep trajectories and mediators of poor sleep: findings from the longitudinal analysis of 41,094 participants of the UK Biobank cohort. Sleep Medicine. 2020;76:120–7.

11. Afolalu EF, Ramlee F, Tang NKY. Effects of sleep changes on pain-related health outcomes in the general population: A systematic review of longitudinal studies with exploratory meta-analysis. Sleep Med Rev. 2018;39:82–97.

12. Amiri S, Behnezhad S. Sleep disturbances and back pain : Systematic review and metaanalysis. Neuropsychiatr. 2020;34(2):74–84.

13. Sivertsen B, Lallukka T, Salo P, et al. Insomnia as a risk factor for ill health: results from the large population-based prospective HUNT Study in Norway. J Sleep Res. 2014;23(2):124–32.

14. Härmä M, Ropponen A, Hakola T, et al. Developing register-based measures for assessment of working time patterns for epidemiologic studies. Scand J Work Environ Health. 2015;41(3):268–79.

15. d’Ettorre G, Vullo A, Pellicani V, et al. Acute low back pain among registered nurses. Organizational implications for practice management. Ann Ig. 2018;30(6):482–9.

16. June KJ, Cho S-H. Low back pain and work-related factors among nurses in intensive care units. Journal of Clinical Nursing. 2011;20(3-4):479–87.

17. Burdelak W, Bukowska A, Krysicka J, et al. Night work and health status of nurses and midwives. cross-sectional study. Med Pr. 2012;63(5):517–29.

18. Matre D, Nilsen KB, Katsifaraki M, et al. Pain complaints are associated with quick returns and insomnia among Norwegian nurses, but do not differ between shift workers and day only workers. Int Arch Occup Environ Health. 2020;93(3):291–9.

19. Yao Y, Zhao S, An Z, et al. The associations of work style and physical exercise with the risk of work-related musculoskeletal disorders in nurses. Int J Occup Med Environ Health. 2019;32(1):15–24.

20. Eriksen W, Bruusgaard D, Knardahl S. Work factors as predictors of intense or disabling low back pain; a prospective study of nurses’ aides. Occup Environ Med. 2004;61(5):398–404.

21. Wang Y, Xie J, Yang F, et al. The prevalence of primary headache disorders and their associated factors among nursing staff in North China. J Headache Pain. 2015;16:4.

22. Bjorvatn B, Pallesen S, Moen BE, et al. Migraine, tension-type headache and medication-overuse headache in a large population of shift working nurses: a cross-sectional study in Norway. BMJ Open. 2018;8(11):e022403.

23. Xie W, Li R, He M, et al. Prevalence and risk factors associated with headache amongst medical staff in South China. J Headache Pain. 2020;21(1):5.

24. Carney CE, Buysse DJ, Ancoli-Israel S, et al. The Consensus Sleep Diary: Standardizing Prospective Sleep Self-Monitoring. Sleep. 2012;35(2):287–302.

25. Hilbe JM. Negative Binomial Regression. 2 ed. Cambridge: Cambridge University Press; 2011.

26. Hayes AF, Rockwood NJ. Regression-based statistical mediation and moderation analysis in clinical research: Observations, recommendations, and implementation. Behaviour Research and Therapy. 2017;98:39–57.

27. Geldhof GJ, Anthony KP, Selig JP, et al. Accommodating binary and count variables in mediation: A case for conditional indirect effects. International Journal of Behavioral Development. 2017;42(2):300–8.

28. Zhang Q, Chair SY, Lo SHS, et al. Association between shift work and obesity among nurses: A systematic review and meta-analysis. Int J Nurs Stud. 2020;112:103757.

29. Zheng H, Chen C. Body mass index and risk of knee osteoarthritis: systematic review and meta-analysis of prospective studies. BMJ Open. 2015;5(12):e007568.

30. Zhang T-T, Liu Z, Liu Y-L, et al. Obesity as a Risk Factor for Low Back Pain: A Meta-Analysis. Clinical Spine Surgery. 2018;31(1):22–7.

31. Ritonja J, Aronson KJ, Matthews RW, et al. Working Time Society consensus statements: Individual differences in shift work tolerance and recommendations for research and practice. Ind Health. 2019;57(2):201–12.

32. Stone AA, Broderick JE. Real-Time Data Collection for Pain: Appraisal and Current Status. Pain Medicine. 2007;8(3):S85–S93.

33. Buckley JP, Keil AP, McGrath LJ, et al. Evolving methods for inference in the presence of healthy worker survivor bias. Epidemiology. 2015;26(2):204–12.

34. Driesen K, Jansen NW, van Amelsvoort LG, et al. The mutual relationship between shift work and depressive complaints--a prospective cohort study. Scand J Work Environ Health. 2011;37(5):402–10.

35. Pieh C, Jank R, Waiß C, et al. Night-shift work increases cold pain perception. Sleep Med. 2018;45:74–9.

36. Takahashi M, Iwakiri K, Sotoyama M, et al. Musculoskeletal pain and night-shift naps in nursing home care workers. Occupational Medicine. 2009;59(3):197–200.

37. Karcioglu O, Topacoglu H, Dikme O, et al. A systematic review of the pain scales in adults: Which to use? The American Journal of Emergency Medicine. 2018;36(4):707–14.

38. Statistics Norway. 12546: Employees with health care education. 4th quarter, by sex, age, contents, year and health care education 2021 [Available from: https://www.ssb.no/en/statbank/table/12546/tableViewLayout1/.

39. Maich KHG, Lachowski AM, Carney CE. Psychometric Properties of the Consensus Sleep Diary in Those With Insomnia Disorder. Behavioral Sleep Medicine. 2018;16(2):117–34.

40. Johnson TJ, Basu S, Pisani BA, et al. Depression predicts repeated heart failure hospitalizations. J Card Fail. 2012;18(3):246–52.

