## Supplementary table 1 for "The association between proportion of night shifts and musculoskeletal pain and headaches in nurses: a cross-sectional study"

**Supplementary table 1.** Estimates for the conditional direct effects and the conditional indirect effects for total sleep time (TST) as a potential mediator for the association between proportion of night shifts and musculoskeletal pain and headaches among nurses (n=645). Presented with incidence rate ratio (IRR) with 95 % confidence intervals (CI).

| | **Participants, n** | **Adjusted IRR (95 % CI)[a]** |
|---|---|---|
| **Proportion of night shifts (%)** | **Neck-, shoulder-, and upper back pain** | |
| Direct effect <25 | 399 | 1.00 (ref.) |
| Direct effect 25-49.9 | 141 | 0.96 (0.75, 1.23) |
| Direct effect ≥50 | 105 | 0.87 (0.66, 1.14) |
| Indirect effect 25-49.9 | 141 | 1.01 (0.99, 1.02) |
| Indirect effect ≥50 | 105 | 1.01 (0.99, 1.04) |
| **Proportion of night shifts (%)** | **Low back pain** | |
| Direct effect <25 | 399 | 1.00 (ref.) |
| Direct effect 25-49.9 | 141 | 0.90 (0.68, 1.20) |
| Direct effect ≥50 | 105 | 0.79 (0.58, 1.09) |
| Indirect effect 25-49.9 | 141 | 1.00 (0.99, 1.01) |
| Indirect effect ≥50 | 105 | 1.00 (0.99, 1.01) |
| **Proportion of night shifts (%)** | **Pain in the upper extremities** | |
| Direct effect <25 | 399 | 1.00 (ref.) |
| Direct effect 25-49.9 | 141 | 0.78 (0.53, 1.16) |
| Direct effect ≥50 | 105 | 0.89 (0.56, 1.37) |
| Indirect effect 25-49.9 | 141 | 1.00 (1.00, 1.00) |
| Indirect effect ≥50 | 105 | 1.00 (1.00, 1.00) |
| **Proportion of night shifts (%)** | **Pain in the lower extremities** | |
| Direct effect <25 | 399 | 1.00 (ref.) |
| Direct effect 25-49.9 | 141 | 0.92 (0.70, 1.22) |
| Direct effect ≥50 | 105 | 0.69 (0.51, 0.95) |
| Indirect effect 25-49.9 | 141 | 1.00 (1.00, 1.01) |
| Indirect effect ≥50 | 105 | 1.00 (1.00, 1.01) |
| **Proportion of night shifts (%)** | **Headache** | |
| Direct effect <25 | 399 | 1.00 (ref.) |
| Direct effect 25-49.9 | 141 | 1.05 (0.86, 1.27) |
| Direct effect ≥50 | 105 | 0.96 (0.77, 1.19) |
| Indirect effect 25-49.9 | 141 | 1.03 (0.96, 1.09) |
| Indirect effect ≥50 | 105 | 1.05 (0.96, 1.15) |

[a]Adjusted for age and BMI
